# Exploring consumer preferences for cannabis edible products to support public health policy: A discrete choice experiment

**DOI:** 10.1101/2023.09.20.23295824

**Authors:** Jennifer R. Donnan, Karissa Johnston, Michael Coombs, Maisam Najafizada, Lisa D. Bishop

**Author notes:** Corresponding Author: Dr. Jennifer Donnan School of Pharmacy, Memorial University of Newfoundland 300 Prince Phillip Drive St. John’s, NL, Canada A1B 3V6.

## Abstract

**Background:** In October 2019, cannabis edibles were legalized for sale in Canada. This move was intended to improve public safety by regulating contents (including a maximum 10 mg tetrahydrocannabinol (THC) per package) and packaging to prevent accidental ingestion or over consumption. This study aimed to explore consumer preferences for cannabis edibles to inform cannabis policy.

**Methods:** We explored the relative importance and trade-offs consumers make for attributes of cannabis edibles using a discrete choice experiment. Attributes included type of edible, price, THC content, cannabis taste, package information, product consistency, product recommendations, and Health Canada regulation. Participants lived in Canada, were 19 years of age or older, and purchased a cannabis edible in the last 12 months. A multinomial logit (MNL) model was used for the base model, and latent class analysis to assess preference sub-groups.

**Results:** Among 684 participants, the MNL model showed that potency was the most relevant followed by edible type. A two-group latent class model revealed two very distinct preference patterns. Preferences for group 1 (∼65% of sample) were driven primarily by edible type, while for group 2 (∼35% of sample) were driven almost entirely by THC potency.

**Conclusion:** This study found that consumer preferences for ∼65% of consumers of cannabis edibles are being met through regulated channels. The remaining ∼35% are driven by THC potency at levels that are not currently available on the licensed market. Attracting this market segment will require reviewing the risks and benefits of restricting THC package content.

## 1. Introduction

On October 17, 2018, Canada became the second country to legalize cannabis, starting with dried flower products. One year later, additional product types including cannabis vapes and edibles were approved for sale (1). Its legalization and regulation changed the way Canadians could access cannabis to promote health and safety. From the 2017 Canadian Cannabis Survey (CCS) (2), 22% of those 16 and older reported using cannabis in the last 12 months, with greater use among those aged 16-24 (41%) compared to those aged 25 years and older (18%). More males (26%) reported past 12-month use versus females (18%). During the year after cannabis was first legalized, 37% of individuals obtained cannabis from a legal storefront or online source (3). A greater– albeit slower– transition to licensed sources was observed in the years to follow. Purchases from legal and licensed sources jumped in 2020 (54%) and expanded further in 2021 (64%) (4). Despite this, unlicensed sources still compose a great portion of sales, reinforcing the need for further efforts and consideration of what consumers value (5).

Smoking cannabis is the most common method of consumption regardless of province or territory; however, the use of other product types is expanding. National survey data demonstrated that the prevalence of edible use has increased since legalization from 32% in 2017 to 53% in 2021 (2,4). Data from the United States has shown that those individuals who consume edibles tend to be heavier cannabis users, with more frequent use and longer periods spent high compared to those who do not consume edibles (6). While edibles have the benefit of not carrying the respiratory health impacts of smoking and vaping, they are not benign with respect to health consequences. More frequent edible cannabis consumption has been significantly associated with physical dependence, impaired control, academic/occupational problems, self-care problems, and risk behavior, after controlling for demographics and socioeconomic characteristics (7). Additionally,due to the delayed effects of edible cannabis, studies have shown edibles to be more likely to result in unexpected highs among adults (8). Unintentional pediatric exposure to cannabis also increased after decriminalization in certain US states. Most of the more serious exposures were a result of ingestion, which was believed to be due to their increased palatability over other cannabis forms as well as the typically higher THC concentrations (9).

These public health and safety considerations were the reason behind Canada’s approach to strictly limit the amount of THC in edible cannabis products. Canadian federal regulations limit the amount of THC to 10 mg per package regardless of the number of edible items in the package (10). Some consumers, in particular those who require higher doses to achieve their desired effect, have stated that Health Canada-approved cannabis is cost prohibitive and too calorically dense at such low doses of THC per package (11). The extent to which limited THC content impacts decisions to purchase from either licensed or unlicensed sources is not clear, nor is the relative impact of other attributes such as package information, taste, or consistency in dose across units.

The multi-attribute utility theory states that when people make decisions, they take into account various attributes of the options presented to them and then make trade-offs between those attributes to optimize personal preferences (12). Discrete choice experiments (DCE) are used to measure the strength of consumer preferences for the attributes of decisions via a survey-based approach. Within these surveys, participants choose between two hypothetical options, each described by a set choice of attributes. Based on the participants’ repeat selections where the hypothetical options are altered slightly within the attributes, the relative importance of each attribute can be quantified. Knowledge of the trade-offs that consumers make for edible cannabis products is key to refining public policy to encourage greater uptake of regulated over unregulated products. The purpose of this study was to quantify consumer preferences for attributes of edible products using a DCE.

## 2. Methods

### 2.1 Study Design

A survey consisting of a four unique DCE questions, including one focused on attributes for cannabis edible products, was used to solicit preferences from cannabis consumers across Canada. This study was carried out following the general framework for good research practices as outlined for conducting DCEs by the International Society for Pharmacoeconomics and Outcomes Research (13). This study is part of a series of studies that explored consumer preferences for different types of cannabis products. Earlier work includes a systematic review of the literature to identify attributes of importance for cannabis consumers (14), focus groups and interviews with cannabis consumers (11) and two DCEs focused on consumer preferences for cannabis vapes (15) and dried flower (16). Detailed methods for the current study have been previously published (15), presented here is a condensed summary.

Data from the systematic review, focus groups and interviews were used to identify attributes and levels that are both important to consumers and policy-relevant for cannabis edible products (Table 1). While we know that price and the amount of THC would be relevant, we also explored the type of edible, cannabis taste, package information, dosing consistency, product recommendations, and if it was regulated by Health Canada. The type of edible refers to the food type (e.g. candy, baked good, savory product) and while not all of these are available in the legal market, they are available through non-licensed channels and impacting consumer choices. Taste gets at preference distinctions between products with a cannabis flavor over a masked flavor (e.g. fruit) Consumers reported that they want access to detailed product information, and not just what is required on a Health Canada approved label, but also information on terpene profiles and cultivation history (11). In previous work, it was found that consistency between servings of homemade edibles (e.g., cookies) was problematic, and accurate knowledge of dose per serving influenced decisions for some consumers (11). The attribute of product recommendations was used to get at the impact of social influences on choices, and what sources of recommendations were most relevant to impact ultimate purchase decisions. Finally, we wanted to include an attribute that explored the impact of having the product regulated by Health Canada, and to see if attributes were more important than Health Canada regulation.

**Table 1:** Attributes and levels for one package of cannabis edibles.

| Attribute | Levels |
| --- | --- |
| **Type of Edible** | A Candy (e.g. chocolate bar, gummy, mint)<br>A Baked Product (e.g. brownie, cookie, granola bar)<br>A Savory Product (e.g. pretzels, trail mix) |
| **Price for Package** | \$5, \$10, \$15 |
| **Amount of TCH per Package** | 5 mg<br>10 mg<br>50 mg<br>100 mg |
| **Cannabis Taste** | Strong cannabis taste<br>Mild cannabis taste<br>No cannabis taste |
| **Package Information** | No info on the package<br>Producer, Amount of THC and/or CBD in milligrams, nutritional information<br>Producer, Amount of THC and/or CBD in milligrams, nutritional information, strain, terpenes, growth and supply Chain Info |
| **Consistency of THC across servings** | Unknown<br>Exactly the same |
| **Product Recommendation** | Recommended by person selling<br>Recommended by family or friend<br>Recommended in online reviews<br>Self-selected without input from others |
| **Regulated by Health Canada** | Yes<br>No<br>Unknow |

The DCE choice task included two unlabeled alternatives, meaning each combination of attribute levels was described as “Option A” or “Option B”, which does not hold any meaning (17) (Figure 1). A fractional factorial design was used. A total of eight choice tasks were included, which allowed for a standard error below the threshold of 0.05. The DCE question was prefaced by a description of a scenario to help frame the choice which the consumer was asked to make. Additional questions on sociodemographic characteristics (e.g. age, province, sex, gender), cannabis consumption, and purchasing history were also included.

**Figure 1.**
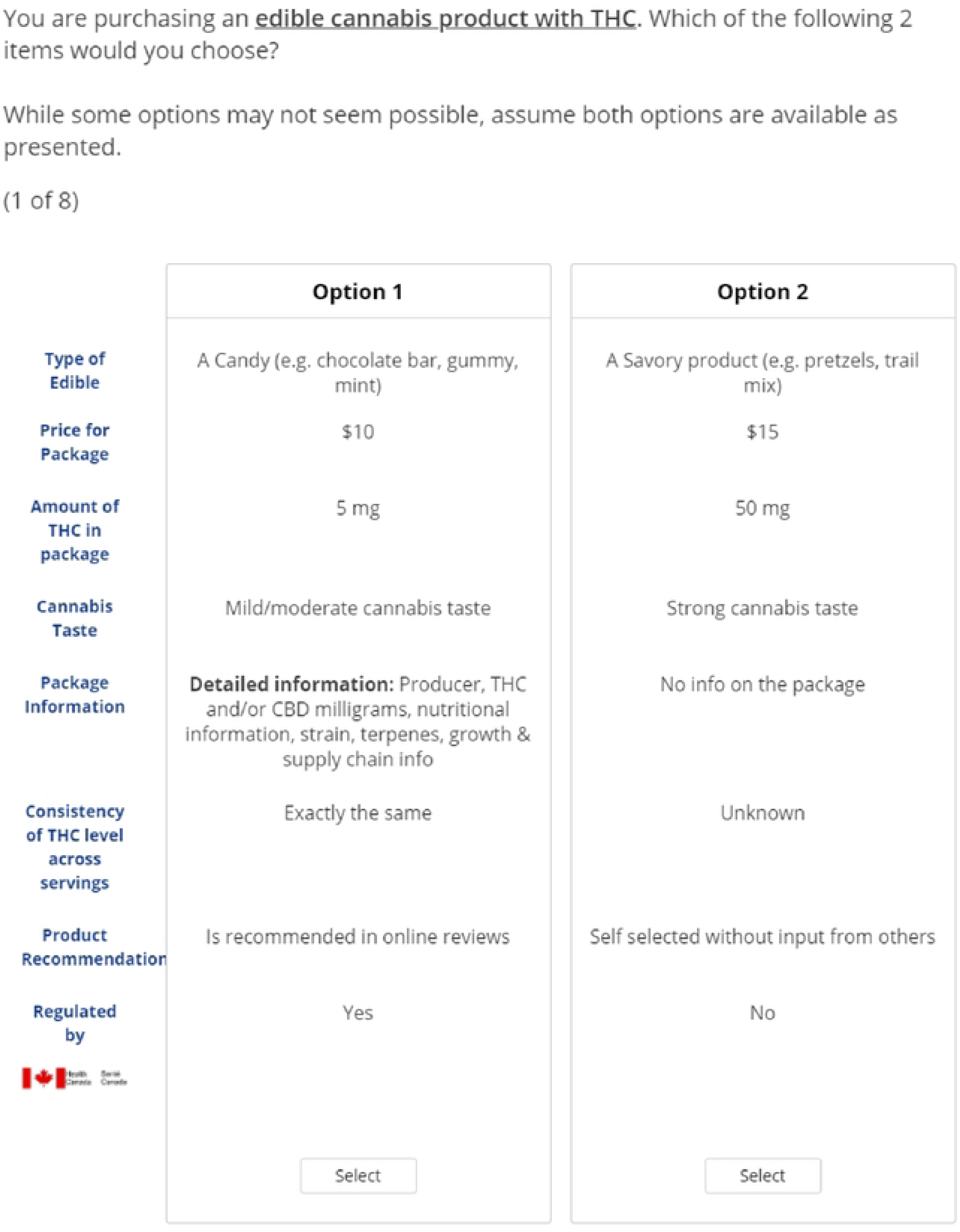
Edible Cannabis Sample Choice Task

### 2.2 Participants

Participants were eligible to complete the survey if they lived in Canada, were 19 years of age or older, and reported having purchased cannabis within the last 12 months. Only those who indicated they had purchased a cannabis edible in the past 12 months were eligible to complete the edible specific DCE. An online research company (Angus Reid) used email solicitation to recruit a representative sample from their proprietary panel between October 8-25^th^, 2021. Interested individuals provided electronic consent within the Sawtooth survey instrument. Only consenting participants proceeds to answer survey questions. Data from respondents who were eligible for the edible DCE and completed the full survey were included in the analysis.

### 2.3 Analysis

Descriptive statistics were used for sample characteristics. Analysis of the DCE data was completed within Sawtooth (Lighthouse Studio) software and included a counts analysis and two regression models, a multinomial logit (MNL) model and a latent class model.

The MNL model was used for the base analysis to calculate average preferences across the sample. The data for each attribute was effects coded except for cost where continuous coding was used to allow for interpretable willingness to pay (WTP) values. Using the least desirable level from each attribute as a reference, odds ratios were calculated. WTP was calculated by estimating the marginal rate of substitution (MRS) by taking the ratio of two co-efficients, with the linear cost estimate used for the comparison attribute.

Finally, a latent class model was used to examine potential sub-groups of preferences within the consumer population. The model of best fit was assessed by selecting the number of latent classes with the lowest CAIC (Consistent Akaike Information Criterion) and BIC (Bayesian Information Criterion) values (18,19). Segment membership probabilities estimated by Sawtooth were used to explore differences in participant characteristics between the groups. Chi-squared tests were used to assess significant differences with key demographic characteristics of the sample (e.g. age, sex, income, province of residence) as well as cannabis use behaviors (e.g. purchase and consumption frequency, reason for use, length of time of use).

### 2.4 Ethical Considerations

This study was carried out in accordance with the Tri-Council Policy Statement and approval by the Memorial University Interdisciplinary Committee on Ethics in Human Research (File #20210143).

## 3. Results

Of the 3,261 individuals who started the survey, of which 1,920 consented and were eligible, and 1626 completed the full survey. The survey consisted of four unique DCE questions and not all participants were eligible for each question. The findings here represent the sample of 684 who completed the DCE focused on edible cannabis purchase decisions. Just over half of the sample identified as men, and about a third were between 30 to 39 years of age. The vast majority (91.8%) had at least some-post secondary education (Table 2).

**Table 2:** Respondent Characteristics.

| Characteristic |  | Number (%)<br>N=684 |
| --- | --- | --- |
| Sex | Female | 333 (48.7) |
|  | Male | 344 (50.3) |
|  | Prefer not to say | 7 (1.0) |
| Gender | Woman | 322 (47.1) |
|  | Man | 343 (50.1) |
|  | Gender Diverse | 8 (1.2) |
|  | Other | 6 (0.9) |
|  | Prefer not to say | 5 (0.7) |
| Age | 19-29 | 146 (21.3) |
|  | 30-39 | 238 (34.8) |
|  | 40-49 | 95 (13.9) |
|  | 50-59 | 94 (13.7) |
|  | 60 or above | 111 (16.2) |
| Race | Black | 12 (1.8) |
|  | East/Southeast Asian | 18 (2.6) |
|  | Latino | 5 (0.7) |
|  | Middle | 7 (1.0) |
|  | South Asian | 14 (2.0) |
|  | White | 628 (91.8) |
|  | Other (please specify): | 26 (3.8) |
| Province | British Columbia | 79 (11.5) |
|  | Alberta | 82 (12.0) |
|  | Saskatchewan | 67 (9.8) |
|  | Manitoba | 65 (9.5) |
|  | Ontario | 115 (16.8) |
|  | Quebec | 41 (6.0) |
|  | New Brunswick | 42 (6.1) |
|  | Nova Scotia | 89 (13.0) |
|  | Prince Edward Island | 18 (2.6) |
|  | Newfoundland and Labrador | 81 (11.8) |
|  | Territories | 5 (0.6) |
| Education | Did not complete high school | 7 (1.0) |
|  | High school diploma | 49 (7.2) |
|  | Some post-secondary | 102 (14.9) |
|  | College/trade/technical/ vocational training completed | 221 (32.3) |
|  | Undergraduate degree | 197 (28.8) |
|  | Graduate degree | 108 (15.8) |
| Employment | Full time student | 65 (9.5) |
|  | Part time student | 18 (2.6) |
|  | Unemployed, but seeking employment | 31 (3.5) |
|  | Unemployed by choice | 8 (1.2) |
|  | Unemployed due to disability | 19 (2.8) |
|  | Employed part time | 57 (8.3) |
|  | Employed full time | 367 (53.7) |
|  | Self employed | 69 (10.1) |
|  | Retired | 90 (13.2) |
|  | Other (please specify:) | 13 (1.9) |
| Income | <\$25,000 | 53 (7.7) |
| | \$25,000 to \$49,999 | 118 (17.3) |
| | \$50,000 to \$74,000 | 122 (17.8) |
| | \$75,000 to \$99,999 | 108 (15.8) |
| | \$100,000 or more | 228 (33.3) |
|  | Prefer not to say | 55 (8.0) |
| Frequency of Cannabis purchase in last 12 months | < 1 per month | 313 (45.8) |
|  | 1-2 times per month | 238 (34.8) |
|  | 3 or more times per month | 133 (19.4) |
| Cannabis consumption frequency | Less than once per month | 132 (19.3) |
|  | At least once per month, less than once per week | 126 (18.4) |
|  | At least once per week | 152 (22.2) |
|  | Once per day | 126 (18.4) |
|  | Multiple times per day | 146 (21.3) |
|  | Prefer not to answer | 2 (0.3) |
| Reason for cannabis use | Medical (Self Prescribed) | 65 (9.5) |
|  | Medical (Authorized) | 26 (3.8) |
|  | Non-medical | 277 (40.6) |
|  | Both medical and non-medical | 307 (44.9) |
|  | Other | 8 (1.2) |
| Initiation of Cannabis Use | Since legalization | 120 (17.5) |
|  | Used in the past then started again since legalization | 252 (36.8) |
|  | Regular user prior to legalization | 312 (45.6) |
| Cannabis Purchase Location | Licensed in-person store | 553 (80.8) |
|  | Licensed online store | 288 (42.1) |
|  | Licensed Medical Dispensary | 66 (9.6) |
|  | Unlicensed in-person store | 87 (12.7) |
|  | Unlicensed online stores | 189 (27.6) |
|  | Unlicensed connection on the community | 160 (23.4) |
|  | Other | 22 (3.2) |

**Table 3:**
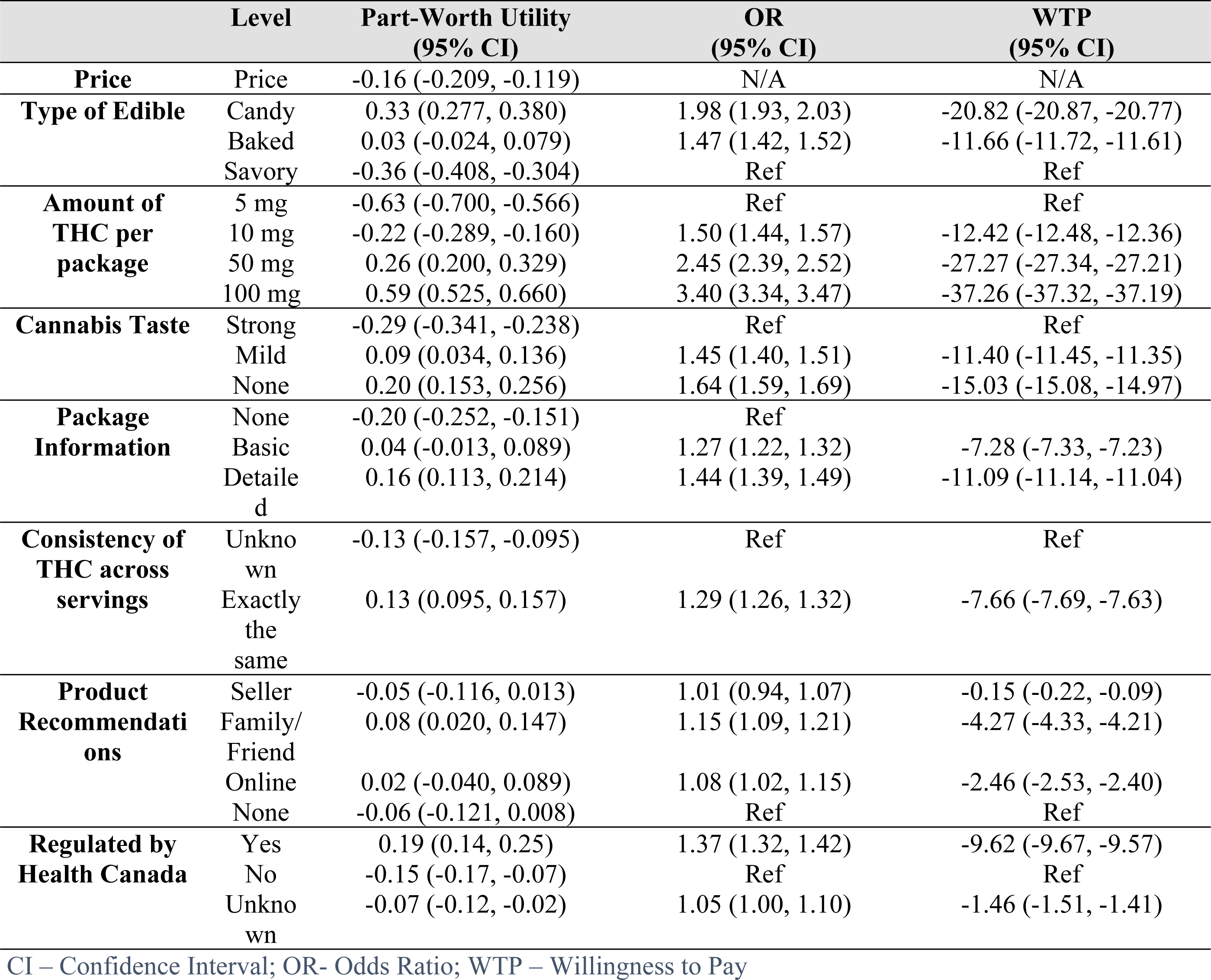
Relative importance of attributes for cannabis edibles using a multinomial logit model.

|  | Level | Part-Worth Utility<br>(95% CI) | OR<br>(95% CI) | WTP<br>(95% CI) |
| --- | --- | --- | --- | --- |
| **Price** | Price | -0.16 (-0.209, -0.119) | N/A | N/A |
| **Type of Edible** | Candy | 0.33 (0.277, 0.380) | 1.98 (1.93, 2.03) | -20.82 (-20.87, -20.77) |
|  | Baked | 0.03 (-0.024, 0.079) | 1.47 (1.42, 1.52) | -11.66 (-11.72, -11.61) |
|  | Savory | -0.36 (-0.408, -0.304) | Ref | Ref |
| **Amount of**<br>**THC per**<br>**package** | 5 mg | -0.63 (-0.700, -0.566) | Ref | Ref |
|  | 10 mg | -0.22 (-0.289, -0.160) | 1.50 (1.44, 1.57) | -12.42 (-12.48, -12.36) |
|  | 50 mg | 0.26 (0.200, 0.329) | 2.45 (2.39, 2.52) | -27.27 (-27.34, -27.21) |
|  | 100 mg | 0.59 (0.525, 0.660) | 3.40 (3.34, 3.47) | -37.26 (-37.32, -37.19) |
| **Cannabis Taste** | Strong | -0.29 (-0.341, -0.238) | Ref | Ref |
|  | Mild | 0.09 (0.034, 0.136) | 1.45 (1.40, 1.51) | -11.40 (-11.45, -11.35) |
|  | None | 0.20 (0.153, 0.256) | 1.64 (1.59, 1.69) | -15.03 (-15.08, -14.97) |
| **Package**<br>**Information** | None | -0.20 (-0.252, -0.151) | Ref |  |
|  | Basic | 0.04 (-0.013, 0.089) | 1.27 (1.22, 1.32) | -7.28 (-7.33, -7.23) |
|  | Detailed | 0.16 (0.113, 0.214) | 1.44 (1.39, 1.49) | -11.09 (-11.14, -11.04) |
| **Consistency of**<br>**THC across**<br>**servings** | Unknown | -0.13 (-0.157, -0.095) | Ref | Ref |
|  | Exactly the same | 0.13 (0.095, 0.157) | 1.29 (1.26, 1.32) | -7.66 (-7.69, -7.63) |
| **Product**<br>**Recommendations** | Seller | -0.05 (-0.116, 0.013) | 1.01 (0.94, 1.07) | -0.15 (-0.22, -0.09) |
|  | Family/Friend | 0.08 (0.020, 0.147) | 1.15 (1.09, 1.21) | -4.27 (-4.33, -4.21) |
|  | Online | 0.02 (-0.040, 0.089) | 1.08 (1.02, 1.15) | -2.46 (-2.53, -2.40) |
|  | None | -0.06 (-0.121, 0.008) | Ref | Ref |
| **Regulated by**<br>**Health Canada** | Yes | 0.19 (0.14, 0.25) | 1.37 (1.32, 1.42) | -9.62 (-9.67, -9.57) |
|  | No | -0.15 (-0.17, -0.07) | Ref | Ref |
|  | Unknown | -0.07 (-0.12, -0.02) | 1.05 (1.00, 1.10) | -1.46 (-1.51, -1.41) |

CI – Confidence Interval; OR- Odds Ratio; WTP – Willingness to Pay

A two-group latent class model demonstrated the best fit (Table 4). In Group 1, which represented almost 65% of the sample, their choices were driven primarily by edible type (candy preferred to baked goods or savory products), followed by taste (preferred less cannabis flavor) and package information (preferred more detail). Of note, price played a very little role in the decisions. In Group 2, representing 35% of the sample, choices were driven almost entirely by the THC potency (preferred 100 mg package over 5 mg package, OR = 304.3), followed by price (Table 5). Participants in this group were willing to pay nearly $42 more for a package with 100 mg over those with 5 mg when all other attributes remained constant. Other attributes played very little role in the choices for this group. Notably, even though Health Canada regulation played a small role in decisions, participants still demonstrated a preference for regulated over non-regulated products. The Venn diagram, set at a 20% inclusion threshold highlights the likelihood of group membership. About 15% (n=103) of the sample have preference tendencies seen in both groups (Figure 2).

**Figure 2:**
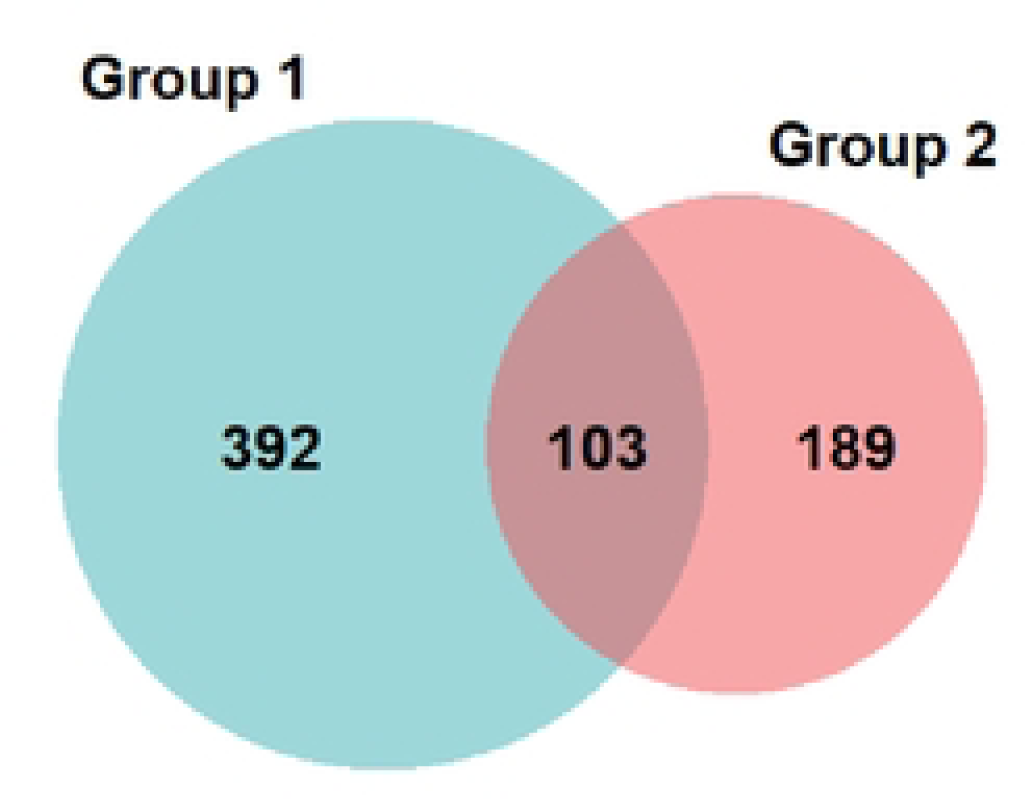
Venn diagram depicting group membership from the latent class model

**Table 4:** Latent class model fit statistics.

| Groups | CAIC | BIC |
| --- | --- | --- |
| 2 | 6319.92 | 6286.92 |
| 3 | 6364.02 | 6314.02 |
| 4 | 6451.71 | 6384.71 |
| 5 | 6548.28 | 6464.28 |

CAIC - Consistent Akaike Information Criterion; BIC - Bayesian Information Criterion

**Table 5:** Relative importance of attributes for cannabis edibles using a latent class model.

|  | Segment Sizes | Part-Worth Utility | OR | WTP | Part-Worth Utility | OR | WTP |
| --- | --- | --- | --- | --- | --- | --- | --- |
|  |  | Group 1 - 65.2% |  |  | Group 2 - 34.8% |  |  |
| **Price for Package** |  | -0.09 | N/A | N/A | -0.68 | N/A | N/A |
| **Type of Edible** | Candy | 0.43 | 2.43 | -48.23 | 0.26 | 1.81 | -4.34 |
|  | Baked | 0.03 | 1.64 | -26.71 | 0.07 | 1.50 | -2.98 |
|  | Savory | -0.46 | Ref | Ref | -0.33 | Ref | Ref |
| **Amount of THC per Package** | 5 mg | -0.19 | Ref | Ref | -2.80 | Ref | Ref |
|  | 10 mg | 0.01 | 1.23 | -11.04 | -1.30 | 4.47 | -10.97 |
|  | 50 mg | 0.10 | 1.34 | -15.91 | 1.17 | 52.98 | -29.08 |
|  | 100 mg | 0.08 | 1.31 | -14.80 | 2.92 | 304.27 | -41.88 |
| **Cannabis Taste** | Strong | -0.35 | Ref | Ref | -0.24 | Ref | Ref |
|  | Mild | 0.10 | 1.58 | -24.66 | 0.03 | 1.31 | -1.97 |
|  | None | 0.25 | 1.82 | -32.59 | 0.21 | 1.56 | -3.28 |
| **Package Information** | None | -0.27 | Ref | Ref | -0.08 | Ref | Ref |
|  | Basic | 0.08 | 1.42 | -19.21 | -0.10 | 0.98 | 0.18 |
|  | Detailed | 0.19 | 1.58 | -24.84 | 0.18 | 1.29 | -1.84 |
| **Consistency of THC across servings** | Unknown | -0.14 | Ref | Ref | -0.19 | Ref | Ref |
|  | Exactly the same | 0.14 | 1.33 | -15.31 | 0.19 | 1.47 | -2.83 |
| **Product Recommendations** | Seller | -0.04 | 1.08 | -3.96 | -0.20 | 0.77 | 1.90 |
|  | Family/Friend | 0.10 | 1.23 | -11.33 | 0.08 | 1.03 | -0.18 |
|  | Online | 0.05 | 1.17 | -4.71 | 0.06 | 1.00 | -1.91 |
|  | None | -0.11 | Ref | Ref | 0.06 | Ref | Ref |
| **Regulated by Health Canada** | Yes | 0.25 | 1.52 | -22.87 | 0.17 | 1.21 | -1.40 |
|  | No | -0.17 | Ref | Ref | -0.02 | Ref | Ref |
|  | Unknown | -0.07 | 1.11 | -5.53 | -0.15 | 0.88 | 0.90 |

OR – Odds Ratio; WTP – Willingness to Pay in Canadian Dollars

The distribution of group membership demonstrated that individuals who were members of group two were significantly more likely to purchase more frequently, consumer more regularly and consumer greater amounts, to consume for recreational purposes and to have consumed cannabis prior to legalization (p-values all <0.001). Age, sex, province or income were not significant predictors of group membership (Table 6).

**Table 6:** Latent class significance of group membership by participant characteristic.

| Factor | Chi-squared | p-value |
| --- | --- | --- |
| Age | 1.65 | 0.800 |
| Sex | 3.03 | 0.219 |
| Province | 19.31 | 0.081 |
| Income | 8.39 | 0.136 |
| Cannabis use in the past 12 months | 35.57 | 0.000 |
| Frequency of cannabis use | 72.09 | 0.000 |
| Amount of cannabis use | 31.82 | 0.000 |
| Purpose of cannabis use | 22.94 | 0.000 |
| Use of cannabis pre-legalization | 41.79 | 0.000 |

## 4. Discussion

This research indicates that the THC content in cannabis edible products plays a major role in Canadian consumers choices to purchase between the licensed and unlicensed markets. This main finding, however, was driven by only a third of the consumer sample population.

Notably, this subset represented a much larger segment of the market, characterized by consumers who purchased more frequently, and consumed more frequently and in larger quantities. These consumers do not have access to the products they seek through the licensed channels. Conversely, approximately 65% of our sample appears to have their preferences met by products available in the licensed market, and this segment of the sample were less concerned with THC potency or price. A report by Deloitte (20) estimated that the market for cannabis alternatives in Canada is valued at $2.7 billion, with about half of this allocated to cannabis edibles. A report using data from the International Cannabis Policy Study survey estimated that only 56% of cannabis edibles are were purchased through legal sources (5).

This is the first study using a DCE to explore consumer preferences for edible cannabis products. In fact, there is very limited evidence on cannabis consumer preferences in general (14), and most studies focused on dried flower as the dominant product type. There is minimal overlap of the relevant attributes between dried flower and edibles and therefore dried flower preference studies cannot be extrapolated to represent such preferences.

While attributes other than THC content did influence purchase choices, factors like edible type, taste, package information, and Health Canada regulation seemed to only influence choices for those whose needs are already met by the licensed market. Shifting more consumers to the licensed market will require changes to regulations limiting THC content, and to a lesser degree price.

In comparison to Canada, regulatory bodies in the United States provide access to higher potency THC edible products. For states that have legalized non-medical cannabis, there is a predetermined standard amount of 5 or 10 mg THC per serving of cannabis edibles. However, packages can contain up to 50 or 100 mg THC in many legalized states and up to 500 mg in the state of Michigan (21,22). Canada’s conservative policy approach to edibles reflects the lack of international experience in codifying laws and the unknown impact on public health and safety. While serious harms are not common with cannabis, edibles pose increased risk due to the delayed onset of effect, increasing consumer risk of overdose (9). For example, there have been case reports of psychosis-related suicide as a result of excessive edible consumption (23). Other research has shown a significant increase in hospitalizations among young children less than 10 years of age (incidence rate ratio 7.49; 95% confidence interval 5.92 – 9.48) due to accidently exposure of cannabis edibles since legalization (24).

The risks of making higher doses of THC available in edible form needs to be weighed against the risks of indirectly encouraging access to such products through unlicensed market. Edible products available on the unlicensed market often contain much higher doses of THC per serving and are not easily distinguished from generic candy or food. Additionally, package labels may not clearly indicate the cannabis contents and the packaging can be made to be more attractive (9), especially to children, often replicating commonly marketed candy. These unregulated products may be more likely to lead to unintentional exposures among adults, children and pets. Any move towards increasing THC potency available in regulated cannabis edibles should be paired with additional safety mechanisms such as restrictions on visually appealing packaging and child friendly flavors (24) and strong public health education campaigns.

Though the amount of THC per package can be much higher in legalized US states, the maximum dosage per serving (referred to as a discrete unit in Canada) is more aligned, with the exception of Michigan. Maximum doses per serving are 10 mg THC in Canada and many US states, although some states limit further to 5 mg per serving. This regulation on serving size ensures a common understanding of the amount of THC per unit, and reduces the chance of accidentally taking larger amounts, and these smaller doses can be easily split for those who seek less than a 10mg dose. Limits to serving size are likely more effective at preventing accidental overdose rather than package limitations (25).

In Canada, a nuanced approach is required to evaluate the risks and benefits of increasing package limits for THC content. To maximize safety, further learnings from jurisdictions in the United States can be explored. For example, one regulatory feature that has been employed in Colorado, Maine, Massachusetts, Nevada is to imprint the THC symbol onto each cannabis edible, making it recognizable when it is out of the package (25). With the cannabis edible market expanding and is subsequent implications for public health and safety, comprehensive public education is also needed to improve public understanding the effects of cannabis edibles, proper storage, and other strategies to protect consumers and prevent accidental exposure.

### 4.1 Limitations

There are several inherent limitations to the discrete choice methodology. These include ordering effect, hypothetical bias and framing effect (13). Strategies to mitigate against these can be found in the supplementary detailed methods. While this study was informed by qualitative data collected from edible cannabis consumers within the Canadian cannabis market the lack of access to higher potency THC products overpowered all other relevant attributes. It would not be fair to say the changes to THC limits alone would shift the bulk of purchases to the licensed market. Replicating this study in the United States where package limits are set to 100 mg (the preferred THC content identified in this current DCE), would help us to understand the attributes of importance in an environment where products available in the licensed market more closely align with those on the unlicensed market. Additionally, product attributes are not the only relevant factors in purchase decisions. Retailer attributes also play a role (11). These could include proximity, customer support, marketing and promotions or availability of product information. Future publications using data from this survey will focus on exploring retailer attributes. Considering these studies together would provide a more complete picture of consumers decision making process. Finally, while every effort was made to capture a representative sample of edible cannabis consumers, the population in the sample does have a higher education and income than the average Canadian population, and predominantly identify as Caucasian. Preferences for people if minority races, or lower socioeconomic status may not be truly reflected in this data.

## 5. Conclusion

This study demonstrated that regulated cannabis edibles are not meeting the needs of about a third of the consumer population; and this segment of the population tends to consist of the more experienced users who purchase and consume cannabis more frequently and in larger quantities. These consumers purchase cannabis on the basis of THC potency and prefer the packages with higher THC content. As a result, these consumers are willing to make trade-offs with purchasing a regulated product to get an unregulated product containing more THC. Although increasing the THC content allowed in each package of cannabis edibles might boost sales of regulated products, the public health implications of such a change remain unclear and warrants further investigation.

## Data Availability

Data can be made available upon request, and with the approval of the Interdisciplinary Committee on Ethics in Human Research

## Acknowledgements

The authors received financial support for conduct of the research from the Canadian Institutes of Health Research (Grant No. RN407334 - 429120) and the Canadian Centre of Substance Use and Addiction for the Partnerships for Cannabis Policy (Grant Nos. RN407334 – 429120 and B2 – RESGRL 413-10-9633) inclusive of this research.

## Author Contributions

**Conceptualization, Methodology:** JD

**Investigation:** JD

**Formal Analysis, Visualization, Writing – Original Draft Preparation**: JD, KJ

**Data curation and Validation:** JD, KJ

**Funding acquisition:** JD, LB, MN

**Resources, software licensing**: JD

**Project Supervision and Administration**: JD

**Writing – Review & Editing:** All authors (JD, LB, MN, MC)

